# INFREQUENT FEEDING MASKING THE BENEFITS OF EXCLUSIVE BREASTFEEDING AT 14WEEKS POST-DELIVERY IN A RURAL SETTING: IMPACT ON INFANT GROWTH AND MATERNAL MENTAL HEALTH

**DOI:** 10.1101/2024.06.07.24308596

**Authors:** Adenike Oluwakemi Ogah

## Abstract

**Background:** Optimal exclusive breastfeeding in terms of frequency of feeds per day and lactational support are often not adequately addressed by healthcare providers. This study examined the relationship between the levels of exclusive breastfeeding, frequency of feeding, alternate feeding patterns, health and growth impacts, among 14 weeks old infants in a remote, understudied village in East Africa.

**Subject and methods:** This was a secondary cross-sectional analysis of a prospective cohort data. Data of 514 out of the 529 mother-newborn pairs recruited from birth were obtained and analysed at 14 weeks post-delivery in the clinic. Infant feeding patterns and growth were assessed. Mothers were interviewed using the Edinburgh postpartum depression score to assess mental status. Parametric, non-parametric and regression analysis were applied to examine the relationship between maternal/infant characteristics, infant feeding frequency/patterns and infant growth. The results were presented in p-values, Odds ratio and 95% confidence interval.

**Results:** 514 infants were examined at the well-infant clinic, 14-weeks post-delivery. Overall, 32 (6.2%), 33 (6.4%), 84 (16.3%) and 87 (16.9%) out of these 514 infants were weight faltering, wasted, stunted and had slow head growth, respectively, based on anthropometry. Only 12 (7.5%) out of these 159 malnourished infants (some infants had >1 type of malnutrition), showed clinical signs of undernourishment. The exclusive breastfeeding (EBF) rate in the infant cohort had dropped further to 83.9% from the previous rates at birth, 6 weeks and 10weeks post-delivery. 106 (20.6%) of these 514 infants were not adequately fed. The complementary feeds offered to the infants in the non-exclusive breastfeeding group included maize/cassava/soya porridge, cow milk and plain water. None was on infant formula. Infrequent feeding was strongly associated with infants in the EBF (OR=2.08; 95% CI 1.04, 4.17; p=0.038) group unlike those in the non-EBF group. Being exclusively breastfed had no significant positive effect on infant growth, probably because it was weakened by infrequent feeding observed in a larger proportion of infants in the EBF group (22.3%) compared to those in the non-EBF group (12.0%). Non-exclusive breastfeeding was significantly associated with increase in maternal depression score (OR 1.2; 95% CI 1.09, 1.32) and overweight infant (OR 1.87; 95% CI 1.02, 3.41). Infrequent feeding was significantly associated with slow head growth (p=0.001), weight faltering (0.021) and wasting (p=0.013). Hence, infants in the EBF group were significantly associated with slow head growth (OR=2.94; 95% CI 1.25, 7.14; p=0.014) and stunting (OR=4.55; 95% CI 1.61, 12.5), probably as a result of infrequent feeding. High maternal depression score was significantly protective against all categories of infant growth faltering (underweight, stunting, wasting and slow head growth). The male infant was 7.66 times (95% CI 4.09, 14.35; p<0.001), 3.28 times (95% CI 1.89, 5.7; p<0.001) and 2.38 times (95% CI 1.07, 5.29; p=0.033) more likely to experience slower head growth, length faltering and weight faltering, respectively, compared to the female infant. Eighty-eight (17.1%) out of the 514 infants had been previously ill before presenting at the clinic; 69 (13.4%) had mild illness i.e. was managed on outpatient-basis; 15 (2.9%) had moderate illness i.e.was hospitalized for ≤7 days; and 4 (0.8%) had severe illness i.e. was hospitalized for >7days. The top 3 most common illnesses observed in these 88 infants were respiratory infections (69.3%), diarrhea (23.9%) and malaria (6.8%).

**Conclusion:** Close attention must be paid to male infants, maternal mental health, type of feeds, frequency of feeding and infant growth parameters during the clinic visits. Infant cow milk feeding practices must be strongly discouraged. The stress of frequent infant feeding and lactational challenges can indeed affect maternal mental health, exclusive breastfeeding practices, and subsequently impact infant growth and health outcomes. Providing comprehensive lactational and mental support programs within maternal and child health systems, can empower mothers to overcome challenges and achieve their exclusive breastfeeding goals.

## Background

The optimal frequency for breastfeeding is typically feeding on demand, which varies from infant to infant but generally ranges from 8 to 12 feeds per day in the first few weeks of life.^1^ Lactational support, such as proper latch methods, timely resolution of breastfeeding issues, and education and encouragement for mothers, plays a critical role in ensuring effective breastfeeding. Exclusive breastfeeding (EBF) has encountered challenges of adherence, despite providing mothers with information about its importance.^2^ A longer duration of exclusive breastfeeding is linked to long-lasting reduced diarrhea-related gut microbiota dysbiosis, along with other benefits.^3,4^

The World Health Organization (WHO) and UNICEF recommend a minimum coverage rate of 90% for exclusive breastfeeding for infants during their first six months of life in poor countries.^5^ It is concerning that, despite the high prevalence of breastfeeding (>95%) in Africa, inappropriate feeding practices such as offering water and other liquids to infants are still common. These practices can undermine exclusive breastfeeding efforts and have negative implications for infant health and nutrition. Despite the promotion efforts by organizations like the WHO and UNICEF, as well as local governments and NGOs, significant challenges persist in achieving and sustaining high rates of exclusive breastfeeding. Additionally, the high rate of bottle-feeding in certain countries (exceeding 30% in Tunisia, Nigeria, Namibia, and Sudan) further exacerbates the issue, as it can contribute to a decline in exclusive breastfeeding rates and potentially lead to health problems for both mothers and infants.^6^

In a systematic review of barriers to exclusive breastfeeding in low and middle-income countries, the authors noted that up to one-third of mothers in Pakistan, Nigeria, and Ghana discontinued exclusive breastfeeding due to factors such as breastfeeding-associated-perceived stress, - frustration and -pain; breast problems, illness, urbanization, and education, which can all contribute to difficulties in maintaining exclusive breastfeeding practices.^7,8^ Additionally, urbanization and higher levels of education among mothers have been linked to shorter durations of breastfeeding, potentially due to factors such as increased reliance on formula feeding, returning to work earlier, or cultural shifts away from breastfeeding. For instance, a study in Ghana documented an exclusive breastfeeding rate at six months as low as 10.3% among city-dwelling professional mothers.^9^

Addressing these challenges requires a multi-faceted approach, which may involve providing lactation support, counseling on breastfeeding techniques, addressing maternal health issues, implementing workplace policies that support breastfeeding mothers, and promoting cultural norms that value and support breastfeeding.

Incomplete exclusive breastfeeding can have far-reaching adverse consequences across various levels of society. Hence, this study was designed to determine the levels of EBF, risk factors, alternative feeding patterns and growth profiles among 514, 14 weeks old infants attending MCH clinic in a rural East African community.

## Materials and methods

The methods employed in carrying out this study is discussed in this section.

### Study setting

Rwanda is a low-income, agricultural and landlocked country with approximately 11 million people living in five provinces, covering an area of 26,338 km.^10^ It is called the home of a ‘1000 hills’.^10^ The limited area of flat land available in most part of Rwanda is a hindrance to farming, animal rearing and construction of standard residential houses, among others.

Rwanda has an average of 4.4 persons per household and a gross domestic product per capita of US $780.80. About half (48%) of its population were under 19 years of age and 39% lived below the poverty line with a life expectancy at birth of 71.1 years for women and an adult literacy rate of 80% among 15–49 years old women. In addition, 87.3% of the population have health insurance and access to health services; spending an average of 47.4 min to reach a health centre.

Gitwe village is located on a high altitude of 1,674 meters above sea level, in the southern province, 240km from Kigali, which is the capital city of Rwanda. Gitwe general hospital began in 1995, immediately after the genocide, for the purposes of providing medical services and later training to this isolated community. The hospital currently has 100% government support, since year 2020. The maximum number of deliveries at the hospital per month was about 200 and about 50 infants visit the maternal and child health clinic (MCH) per day. There are only 2 immunisation days per week.

Some of the challenges in the hospital include poor specialist coverage and few trained health workers, poor supply of equipment, water, electricity, laboratory services and medicines. Challenging cases are referred to the University of Rwanda Teaching Hospital in Butare or Kigali. Gitwe village was selected for this study, because there was no published birth or infant data from this poorly researched, remote, relatively unaccessible community. In 2019, birth and up to 9 months of age feeding and growth data on 529 mother-singleton newborn pairs were compiled in this village, over a period of 12 months in the delivery, postnatal wards and clinics of Gitwe general hospital and at its annex, the MCH.^10^

### Data source and sample

This was a secondary analysis of data collected for a prospective cohort study on growth faltering over a period of 12 months, until infant was 9 months old. Sample size of 529 was calculated using the Epitool sample size calculator.^11^ Mother-newborn pairs were recruited consecutively, on first-come-first-serve basis at birth. Maternal file review and newborn anthropometry [weight (kg), length (cm) and head circumference (cm) measurements, their z-scores and percentiles, were recorded to the nearest decimals] were carried out, soon after birth. Newborn gestational age was determined using the record of the maternal first date of the last menstrual period (LMP), fetal ultrasound dating and/or expanded new Ballard criteria. Weight faltering at 14 weeks post-delivery was defined by NICE static weight assessment of weight-for-age percentile of ≤2.^12^ Weight-for-age, weight-for-length, length-for-age and head circumference-for-age z-scores and percentiles, at 14 weeks were obtained from PediTool calculator, which was based on WHO growth standards and utilised the infants’ corrected age.^13^ Mothers were interviewed using standardized infant feeding questionnaires^14^ and the Edinburgh postpartum depression questionnaire/score^15^ at 14 weeks post-partum in the postnatal clinic. The questionnaires were read to the mothers and filled by the research assistants.

To ensure the quality of data collected, 2 registered nurses were trained as research assistants at Gitwe Hospital on the over-all procedure of mother and newborn anthropometry and data collection by the investigator. The questionnaires were pre-tested before the main data collection phase, on 10 mother-infant pair participants (2% of the total sample). The investigator closely followed the day-to-day data collection process, to ensure completeness and consistency of the questionnaires administered each day, before data entry.

### Statistical analysis

Data clean up, cross-checking and coding were done before analysis. These data were entered into Microsoft Excel statistical software for storage and then exported to Statisty-free web-based online statistics app^16^ for further analysis. Both descriptive and analytical statistical procedures were utilized. Participants’ categorical characteristics were summarised in frequencies and percentages. Chi test was used to determine association between mother/infant characteristics and growth faltering. Numerical characteristics such as maternal depression scores were presented in median and interquartile ranges for skewed data. Non-parametric (for skewed data) and parametric (for normally distributed data) tests were performed to determine relationship between variables and infant growth faltering. Regression models were created to identify the predictors of infant growth faltering and to generate the odds ratio. Factors with p-values <0.1 were included in the regression model. Odds ratio (OR), with a 95% confidence interval (CI) were computed to assess the strength of association between independent and dependent variables. For all, statistical significance was declared at p-value <0.05. The reporting in this study were guided by the STROBE guidelines for observational studies.^17^

### Ethics

The Health Sciences Research Ethics Committee of the University of the Free State in South Africa gave the ethical approval for the collection of the primary data for the original study-’Growth and growth faltering in a birth cohort in rural Rwanda: a longitudinal study’ with Ethical Clearance Number: UFS-HSD2018/1493/2901. Written permission to collect data was obtained from the Director of Gitwe Hospital, Rwanda. Verbal consent were obtained from eligible mothers as they were recruited consecutively from the postnatal ward after delivery, with the attending nurse as a witness. The content of the research information booklet and consent letter were read out to the mothers and their permission was sought after ensuring that they understood the purposes, methods, pros and cons of the research. The participants were given research identity numbers and the principal investigator was responsible for the safe keeping of the completed questionnaires and collected data, to ensure anonymity and confidentiality of the participants.

## Results

The following are the results obtained from the study.

### Participants

Five hundred and ninety-seven (597) babies were delivered at Gitwe Hospital, Rwanda, between 22^nd^ January and 9^th^ May 2019, out of which, eligible 529 mother-newborn pairs were enrolled into the prospective cohort study from birth, Figure 1. 512 of these 529 babies were assessed at 6 weeks postnatal age at the Maternal & Child Health Clinic between 5th March 2019 and 20th June 2019. Between 2^nd^ of April and 18^th^ July 2019, 467 babies from this cohort were reviewed again at 10weeks post-delivery. 514 infants were presented at the 14 weeks-post-delivery clinic between 11^th^ May to 15^th^September 2019.

**Fig 1:**
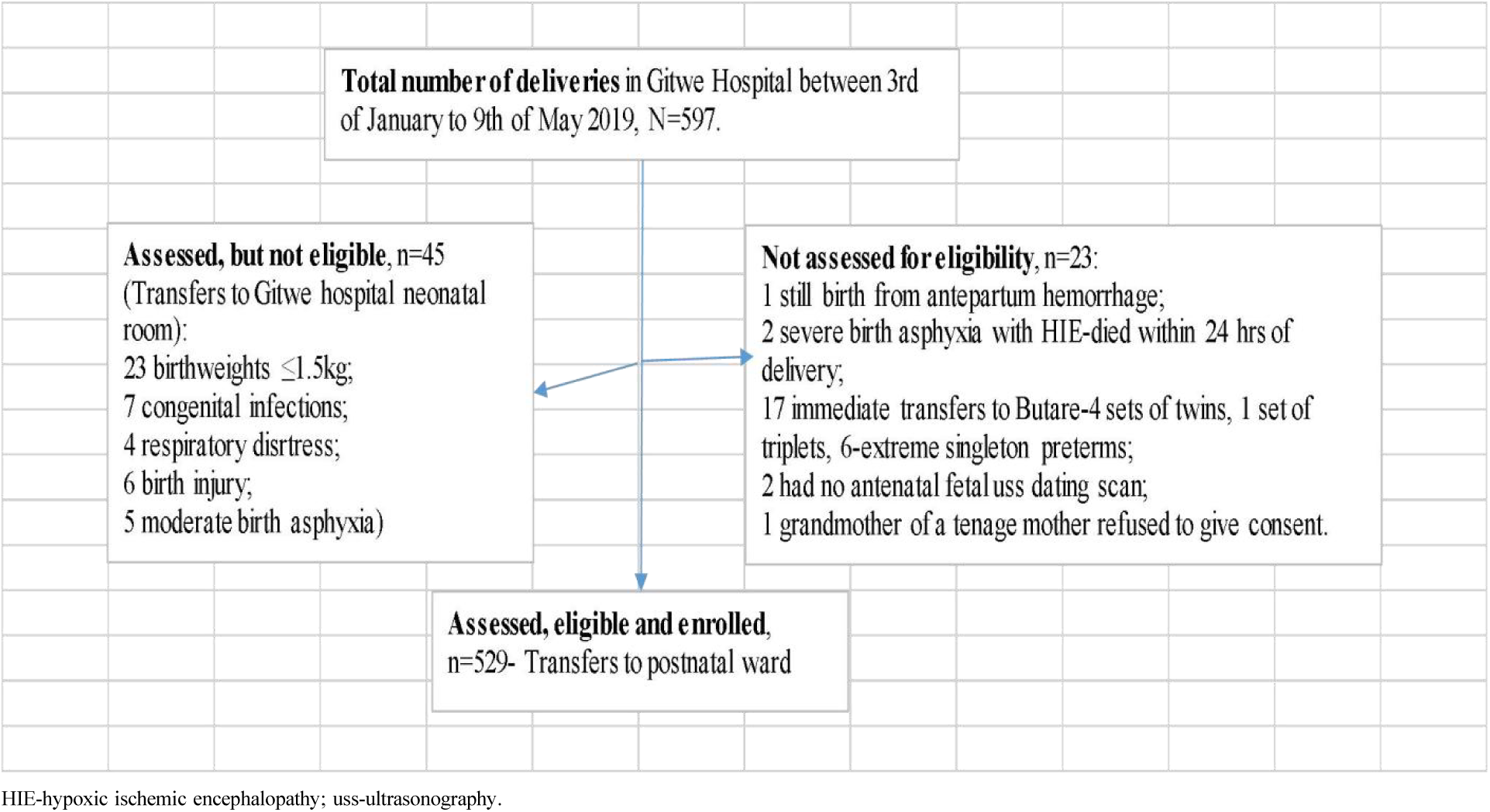
Flow of participants from admission to recruitment into study. Growth and feeding data were collected from a total of 514 infants at 14 weeks post-delivery. 15 out of the 529 (2.8%) babies recruited from birth were absent, hence, 514 infant data at the 14weeks clinic were analysed: 246 girls (47.9%) and 268 boys (52.1%). Their mean age at the 14week-clinic was 3.57 months (sd 0.18).

### Analysis of EBF versus infant and maternal characteristics

The exclusive breastfeeding rate in the infant cohort had dropped further to 83.9% from the previous rates at birth,^18,19^ 6 weeks^20^ and 10weeks^21^ post-delivery. 83 (16.1%) and 106 (20.6%) of these 514 infants were no longer on exclusive breastfeeding and not adequately fed, respectively. The complementary feeds offered to the infants in the non-exclusive breastfeeding group included maize/cassava/soya porridge, cow milk and plain water. None was on infant formula.

#### Infant characteristics and EBF

A higher percentage of girls (17.9%) compared to boys (14.6%); adequately fed (17.9%) infants compared to poorly fed ones (9.4%), p=0.035; those thriving (16.8%) compared to those weight faltering (6.3%); those with normal head growth (18%) compared to those with slow head growth (6.9%), p=0.01; those not stunted (18.4%) compared with those stunted (4.8%), p=0.002; those who were not previously ill (17.4%) compared to those who were ill (10.2%) belonged to the ‘non-exclusive breastfeeding’ group, in Table 1. Stunting (OR=4.55; 95% CI 1.61, 12.5) and slow head growth (OR=2.94; 95% CI 1.25, 7.14; p=0.014) were significantly associated with the infants in the EBF group, Table 1.

**Table 1:**
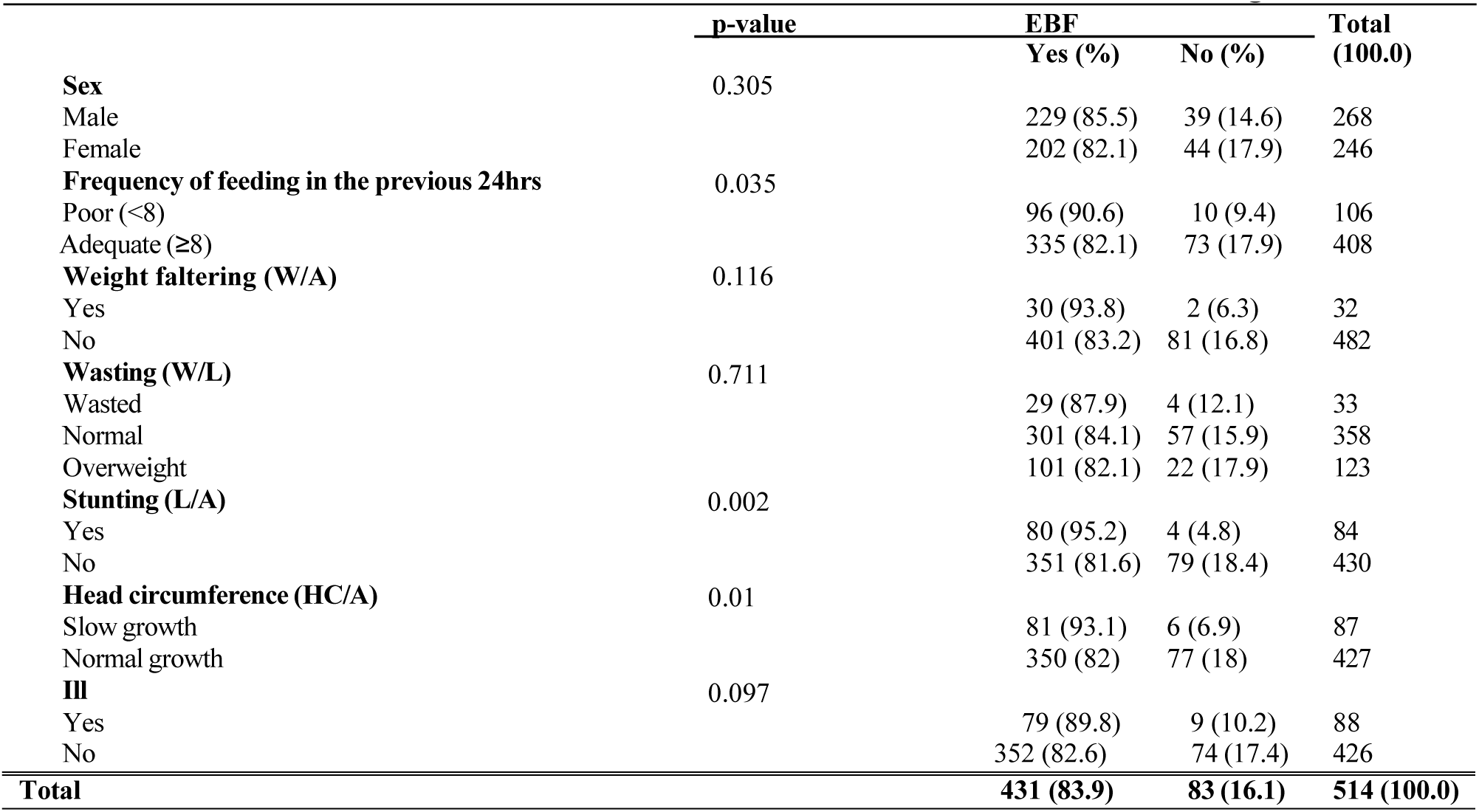
Cross-tabulation of Infants’ characteristics at 14weeks and Exclusive Breastfeeding, n=514.

#### Frequency of feeding in 24hrs

The average frequency of feeding in the exclusively breastfed infants (9.39, sd 2.38; 95% CI 9.29, 9.70) was significantly lower than the average in the non-exclusive breastfeeding group (10.01, sd 2.1), t-test = -2.4 (p=0.018). Poor frequency of feeding was strongly associated with EBF (OR=2.08; 95% CI 1.04, 4.17; p=0.038). Of note, there was no significant difference (p=0.087) in the average frequency of breastfeeding between the infant girl (mean frequency in 24hrs was 9.31, sd 2.34) and boy (mean frequency in 24hrs was 9.66, sd 2.34).

#### Head circumference z-scores

Infants in the EBF group had lower values for the head circumference z-scores (Median = -1.16) than those in the non-EBF group (Median = -0.84). A Mann-Whitney U test was conducted to compare head circumference z-scores between the EBF and non-EBF groups. The test showed that the difference between the 2 groups was statistically significant, U = 13843.5, n1 = 431, n2 = 83 p = .001. The effect size r =0.14 was small.

#### Maternal post-partum depression scores

The mothers in the exclusive breastfeeding group had lower values for postpartum depression (Median = 7, IQR 0,8) than those who were not breastfeeding exclusively (Median = 8, IQR 7.5, 9). A Mann-Whitney U test was conducted to compare the depression scores between the 2 groups of mothers. The difference between those exclusively breastfeeding and those not, with respect to postpartum depression was statistically significant, U = 12193, n1 = 431, n2 = 83 p = <0.001. However, the effect size r was 0.21, which was small.

### Logistic regression analysis of relationship between non-exclusive breastfeeding and infant/maternal characteristics

The logistic regression model was only significant for maternal depression score and overweight infant, where, non-exclusive breastfeeding was associated with increase in maternal depression score (OR 1.2; 95% CI 1.09, 1.32) and overweight infant (OR 1.87; 95% CI 1.02, 3.41), Table 2 .

**Table 2:** Logistic regression analysis between non-exclusive breastfeeding and infant/maternal characteristics.

|  | Coefficient B | Standard error | z | p | Odds Ratio | 95% conf. interval |
| --- | --- | --- | --- | --- | --- | --- |
| Constant | -1.6 | 0.71 | 2.26 | .024 | 0.2 | 0.05 - 0.81 |
| Male infant | -0.17 | 0.26 | 0.66 | .512 | 0.84 | 0.51 - 1.4 |
| Previously ill | -0.44 | 0.41 | 1.08 | .281 | 0.64 | 0.29 - 1.44 |
| Frequency of feeding | -0.11 | 0.08 | 1.35 | .177 | 0.9 | 0.77 - 1.05 |
| Maternal depression score | 0.18 | 0.05 | 3.74 | <.001 | 1.2 | 1.09 - 1.32 |
| Underweight | 0.2 | 0.87 | 0.24 | .814 | 1.23 | 0.22 - 6.75 |
| Overweight | 0.62 | 0.31 | 2.03 | .043 | 1.87 | 1.02 - 3.41 |
| Wasting | 0.34 | 0.67 | 0.52 | .604 | 1.41 | 0.38 - 5.21 |
| Stunting | -1.13 | 0.59 | 1.9 | .057 | 0.32 | 0.1 - 1.03 |
| Slow head growth | -0.56 | 0.48 | 1.17 | .243 | 0.57 | 0.22 - 1.47 |

### Logistic regression analysis of relationship between frequency of feeding and infant/maternal characteristics

The logistic regression model was only significant for maternal depression score, where, poor feeding was associated with lowering of maternal depression score (OR 0.68; 95% CI 0.62, 0.75), Table 3.

**Table 3:** Logistic regression model of frequency of feeding and infant/maternal characteristics.

|  | Coefficient B | Standard error | p | Odds Ratio | 95% conf. interval |
| --- | --- | --- | --- | --- | --- |
| Constant | -0.15 | 0.27 | .573 | 0.86 | 0.5 - 1.46 |
| Male gender | -0.39 | 0.3 | .192 | 0.68 | 0.38 - 1.21 |
| Non-EBF | 0.33 | 0.47 | .481 | 1.39 | 0.55 - 3.5 |
| Previously ill | 0.47 | 0.31 | .125 | 1.6 | 0.88 - 2.91 |
| Maternal depression score | -0.39 | 0.05 | <.001 | 0.68 | 0.62 - 0.75 |
| Underweight | 0.36 | 0.46 | .441 | 1.43 | 0.58 - 3.56 |
| Obese | 0.13 | 0.34 | .699 | 1.14 | 0.59 - 2.2 |
| Wasting | 0.53 | 0.46 | .246 | 1.7 | 0.69 - 4.15 |
| Stunting | 0.18 | 0.37 | .628 | 1.2 | 0.58 - 2.48 |
| Slow head growth | 0.46 | 0.36 | .198 | 1.58 | 0.79 - 3.17 |

### Infant growth at 14 weeks post-delivery

Overall, 32 (6.2%), 33 (6.4%), 84 (16.3%) and 87 (16.9%) out of the 514 infants were weight faltering, wasted, stunted and had slow head growth, respectively. Only 12 (7.5%) infants out of these 159 malnourished infants showed clinical signs of undernourishment. Of note some infants had more than one type of malnutrition.

The male infant was 7.66 times (95% CI 4.09, 14.35; p<0.001), 3.28 times (95% CI 1.89, 5.7; p<0.001) 2.38 times (95% CI 1.07, 5.29; p=0.033) more likely to experience slower head growth, length faltering and weight faltering, respectively, compared to the female infant, Tables 4, 5 and 7. The higher the maternal depression score (OR=0.75; 95% CI 0.63, 0.9; p=0.002) and frequency of feeding (OR=0.78; 95% CI 0.63, 0.96; p=0.021), the less likely the infant will experience weight faltering, Table 4.

**Table 4:**
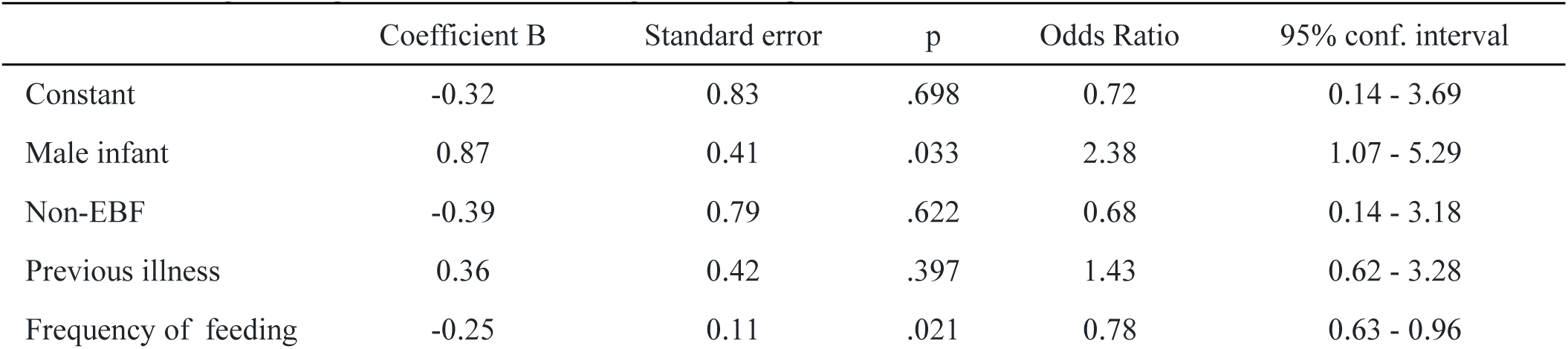
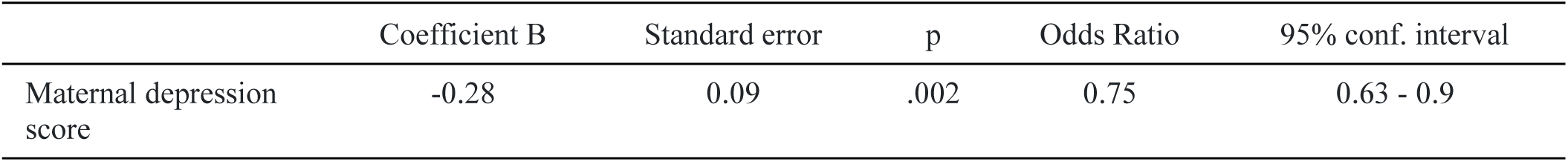
Logistic regression model of weight faltering and infant/maternal characteristics.

|  | Coefficient B | Standard error | p | Odds Ratio | 95% conf. interval |
| --- | --- | --- | --- | --- | --- |
| Constant | -0.32 | 0.83 | .698 | 0.72 | 0.14 - 3.69 |
| Male infant | 0.87 | 0.41 | .033 | 2.38 | 1.07 - 5.29 |
| Non-EBF | -0.39 | 0.79 | .622 | 0.68 | 0.14 - 3.18 |
| Previous illness | 0.36 | 0.42 | .397 | 1.43 | 0.62 - 3.28 |
| Frequency of feeding | -0.25 | 0.11 | .021 | 0.78 | 0.63 - 0.96 |
| Maternal depression score | -0.28 | 0.09 | .002 | 0.75 | 0.63 - 0.9 |

**Table 5:** Logistic regression model of length faltering and infant/maternal characteristics.

|  | Coefficient B | Standard error | z | p | Odds Ratio | 95% conf. interval |
| --- | --- | --- | --- | --- | --- | --- |
| Constant | -0.59 | 0.57 | 1.05 | .295 | 0.55 | 0.18 - 1.67 |
| Male infant | 1.19 | 0.28 | 4.21 | <.001 | 3.28 | 1.89 - 5.7 |
| Non-EBF group | -0.95 | 0.56 | 1.69 | .091 | 0.39 | 0.13 - 1.16 |
| Previous illness | 0.21 | 0.32 | 0.66 | .51 | 1.23 | 0.66 - 2.29 |
| Frequency of feeding | -0.08 | 0.07 | 1.12 | .264 | 0.93 | 0.81 - 1.06 |
| Maternal depression score | -0.24 | 0.05 | 5.22 | <.001 | 0.78 | 0.71 - 0.86 |

The higher the maternal depression score, the less likely the infant will experience length faltering (OR=0.89; 95% CI 0.82, 0.97; p=0.009), Table 5.

The higher the maternal depression score (OR=0.8; 95% CI 0.68, 0.94; p=0.007) and frequency of feeding (OR=0.76; 95% CI 0.62, 0.94; p=0.013), the less likely the infant will experience wasting, Table 6.

**Table 6:** Logistic regression model of wasting and infant/maternal characteristics.

|  | Coefficient B | Standard error | p | Odds Ratio | 95% conf. interval |
| --- | --- | --- | --- | --- | --- |
| Constant | 0.4 | 0.8 | .62 | 1.49 | 0.31 - 7.18 |
| Male infant | -0.72 | 0.4 | .075 | 0.49 | 0.22 - 1.07 |
| Non-EBF group | 0.23 | 0.64 | .715 | 1.26 | 0.36 - 4.39 |
| Previous illness | 0.62 | 0.41 | .128 | 1.85 | 0.84 - 4.1 |
| Frequency of feeding | -0.27 | 0.11 | .013 | 0.76 | 0.62 - 0.94 |
| Maternal depression score | -0.22 | 0.08 | .007 | 0.8 | 0.68 - 0.94 |

The higher the maternal depression score, the less likely the infant will experience slow head growth (OR=0.89; 95% CI 0.82, 0.97; p=0.009), Table 7.

**Table 7:**
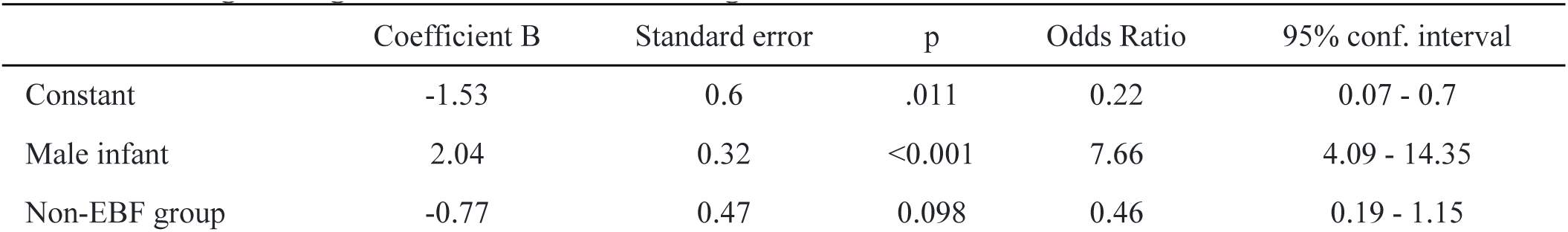
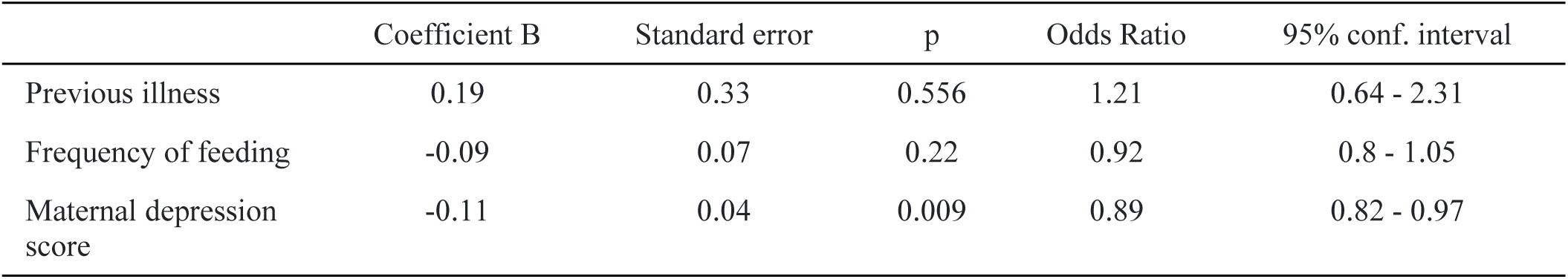
Logistic regression model of slow head growth and infant/maternal characteristics.

When frequency of feeding is at 9times per day, all the growth parameters were within ±1 z-score, Table 8.

**Table 8:**
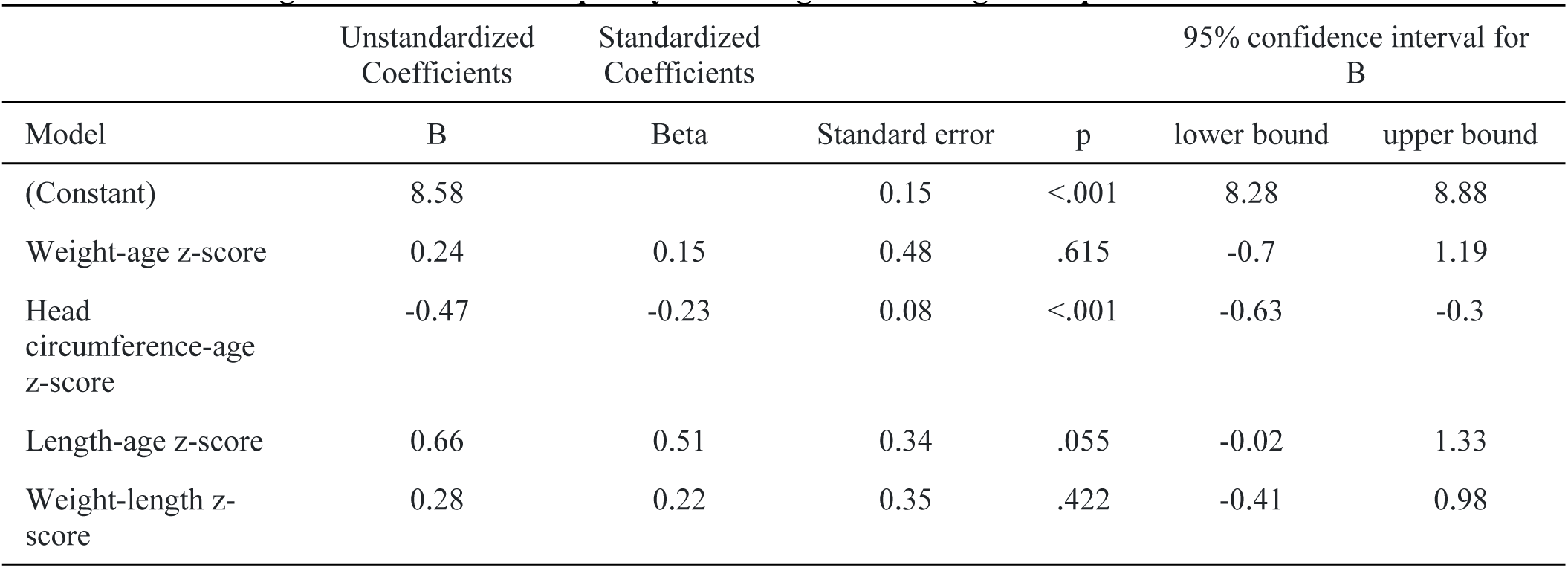
Linear regression model of frequency of feeding and infant growth parameters.

The headcircumference z-score, followed by the length z-score were the most affected of the infants’ growth parameters, Table 9. The ANOVA test showed that each of these growth parameters were significantly different from one another, p<0.001. Hence, each growth parameter should be measured and assessed in each clinic visit.

**Table 9:**
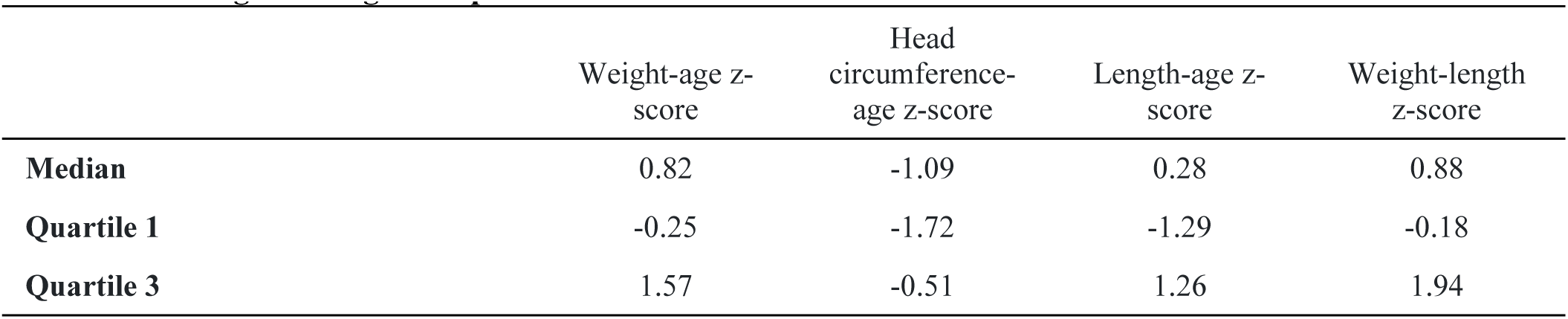
Average infant growth parameters.

|  | Weight-age z-score | Head circumference-age z-score | Length-age z-score | Weight-length z-score |
| --- | --- | --- | --- | --- |
| **Median** | 0.82 | -1.09 | 0.28 | 0.88 |
| **Quartile 1** | -0.25 | -1.72 | -1.29 | -0.18 |
| **Quartile 3** | 1.57 | -0.51 | 1.26 | 1.94 |

### Illness

Eighty-eight (17.1%) out of the 514 infants had been previously ill before presenting at the clinic; 69 (13.4%) had mild illness i.e. was managed on outpatient-basis; 15 (2.9%) had moderate illness i.e.was hospitalised for ≤7 days; and 4 (0.8%) had severe illness i.e. was hospitalised for >7days. Some had more than one type of diagnosis. 61 (69.3%) out of these 88 infants had respiratory problems, 21(23.9%) had diarrhea illness, 6(6.8%) had malaria, 1 (1.1%) had sepsis, 1 (1.1%) had chicken pox and another 1(1.1%) had eye infection. One of the babies with respiratory infection had fracture of the humerus, and another 3 of those with respiratory illness had diarrhea. Of note, the 88 infants, who were previously ill, had significantly lower mean frequency of feeding (8.43, sd 2.71), compared to the 426 infants that were not previously ill (9.71, sd 2.21) 24hrs prior to the 14-week clinic.

## Discussion

This study documented the prevalences of exclusive breastfeeding and infant growth (weight, length, head circumference and weight-length) faltering among 514 infants in the cohort, that presented at the 14th week MCH clinic. In addition, some risk factors associated with infant growth faltering, frequency of feeding and EBF were identified.

Of note, there was a slight improvement in the clinic attendance, from 529 at birth^19,22,23^, to 519 at 6th week,^20^ 467 at the 10th week^21^and then 514 at 14^th^ week clinic. More efforts to improve mother-infant attendance at the post-delivery clinics are needed, as important medical issues in both the infant and the mother may be neglected during this vital period of postnatal life.^24^

All the 514 babies were breastfeeding. Exclusive breastfeeding rate had declined from 96.4% at 6 weeks,^20^ 96.3% at 10weeks^21^and now to 83.9% at 14 weeks postnatal for the same cohort. Though declining, these exclusive breastfeeding rates were still significantly higher than the 59.2% average recorded for infants 0-5months of age in the East African region, according to the 2022 Global Nutrition report.^1^ As the EBF rate dropped, the cohort’s malnutrition rate increased. Mothers who practiced EBF were more likely to feed their infants infrequently, whereas those who did not practice EBF were more likely to feed more frequently, though with less nutritious food, implying that breastfeeding challenges may have led mothers to seek less stressful alternatives to feeding their infants, necessitating immediate investigation and intervention.

The complementary feeds used were mainly carbohydrates and had lower nutritional value than breastmilk, and they were strongly associated with overweight infants and raised maternal depression scores. The cohort’s high infrequent feeding rate obscured the benefits of exclusive breastfeeding (EBF). Notably, the EBF rate fell dramatically from its prior level at a period when working mothers were scheduled to return to work, potentially contributing to infrequent feeding in the EBF group.

Similar observations were made by Al-Katufi et al. in an analytic cross-sectional study^17,18^involving 280 mothers in Saudi Arabia, where only 50% of these mothers were still practicing exclusive breastfeeding after returning to work. Early return to work, deficient breastfeeding support at work, insufficient breast milk and lack of time, as well as inadequate nursing breaks, lactation places, and expressed milk storing facilities inside their workplaces, were identified as major work-related barriers to the continuity of EBF. Maternal mental health was found to influence all areas of infant growth and feeding in this Rwanda study.

Only a small portion (7.5%) of the infants with confirmed undernutrition by anthropometric assessment, could be identified clinically. This emphasizes the importance of regular and comprehensive anthropometry in health facilities. The significant factors in this study promoting infant growth faltering, despite a moderately high exclusive breastfeeding rate, were male gender, low maternal postpartum depression score, and infrequent feeding. As noted during the analysis, the observed sex-based growth faltering was not related to differences in the frequency of feeding, nor to prior illness.

In a systematic review involving meta-analysis of studies conducted in 30 countries globally in 2020, boys were reported to have a higher tendency toward undernutrition, being more prone to wasting, underweight, and stunting compared to girls in 59 out of the 74 studies investigated. This aligns with the findings of this study, although in this study, girls were more prone to wasting; however, this was not statistically significant. The authors have observed that these sex differences in undernutrition are more pronounced during the first 30 months of life and then disappear thereafter, particularly in older age groups. This was attributed to both biologic (immune and hormonal theories) and social factors.^25^

In order to maintain the infant’s growth parameters within ±1 z-score (considered normal) at 14 weeks post-delivery, the average frequency of daily infant feeding should be 9 times. However, according to mothers’ reports, the feeding frequency was as low as 3 times in 24 hours in this study. It is worth noting that no infant was fed with formula, which reflects the very low socio-economic status of the mothers in this cohort.

This observation of low socio-economic status was further supported by a similar cross-sectional study conducted in five health facilities in Kigali, the capital city of Rwanda, in 2021, involving 221 mothers and infants. In that study, 47.1% of mothers offered infant formula, 20.4% gave cow’s milk, and 14.5% gave porridge, among other supplementary feeds. Rates of exclusive breastfeeding (EBF) were much lower in the Kigali urban study (85.1% at birth, 81.9% at 3 months, and 57.5% at 5 months of age) compared to this rural study (94% at birth, 96.4% at 6 weeks, and 96.3% at 10 weeks), suggesting higher levels of socio-economic status among mothers from the Kigali study compared to those in the rural area. In this Kigali study, mothers were observed to commence supplementary feeds as early as 1 month of infant age.^24^

The two studies conducted in Rwanda indicate the widespread practice of feeding infants with cow milk in both urban and rural areas of the country. It is important to note that in rural areas, economic constraints often compel mothers to feed their infants unmodified cow milk. Extensive research has established the negative effects of cow milk on infant health, including causing iron deficiency anemia through gastrointestinal bleeding, inhibiting the absorption of non-heme iron due to its high calcium and casein content, and being a poor source of iron itself. Iron is a crucial nutrient for optimal neurodevelopment in the first year of life.^26^The proportion of growth faltering among infants at 14 weeks post-delivery was observed to be 30.9% overall, with 6.2% experiencing weight faltering in this rural study. This was more than twice as high as the 3.1% weight faltering rate at 6 and 10 weeks and the 3.7% rate at 14 weeks in the same cohort.^20,21^ However, these figures remained notably lower than the 50.4% reported among 4-month-old infants in an urban settings of Sri Lanka, where the exclusive breastfeeding rate at 4 months was 88%.^26,27^

The exclusive breastfeeding rate in Sri Lanka at 4 months exceeded the rate of 83.9% at 14 weeks observed in this Rwanda study. Despite the higher rate of exclusive breastfeeding (EBF) in Sri Lanka, the rate of infant underweight in the country was higher than that in Rwanda. This suggests that factors other than low EBF rate may be contributing to infant malnutrition in Sri Lanka. This discrepancy highlights the need for urgent intervention to address the fast-declining rate of exclusive breastfeeding in this rural, socio-economically deprived community. Among the infant parameters measured in this study, the head circumference exhibited the slowest growth, followed by the length. This suggests the importance of incorporating routine measurement of infant length and head circumference into clinic growth assessments and intensifying the promotion of exclusive breastfeeding in this community. Currently, infant weight and occasionally length are routinely monitored in the Maternal and Child Health (MCH) facilities, while head circumference measurement is seldom performed in these well-child clinics.^26^

It was also noted that adequate infant feeding and thriving were associated with higher maternal depression scores compared to those experiencing growth faltering and poor feeding. This suggests that there may be stress-related experiences associated with breastfeeding and infant care, as indicated by previous studies.^19–22^ Rana et al. also suggested that breastfeeding might be linked to certain mental and physical health issues in these mothers,^7^that could have a negative impact on exclusive breastfeeding and infant care practices.

A strong correlation exists between stressful life events, which may encompass challenges with lactation and infant care, and maternal depression. It seems that these mothers were becoming overwhelmed by breastfeeding challenges at 14 weeks post-delivery, leading them to consider alternatives to exclusive breastfeeding (EBF). This emphasizes the need for closer attention by attending health workers to these mothers, particularly during the first 6 months postpartum, in order to support them in achieving full exclusive breastfeeding and strengthen lactational and mental support in rural health facilities.^28,29^

Although these infants had recovered from their illnesses by the time of the clinic visit, their frequency of feeding in the previous 24 hours was significantly lower than those who had not experienced any ill-health between the 10th and 14th week visits. This may indicate a prolonged period of recuperation from illness that negatively impacted their feeding habits and, consequently, their growth parameters. Acute bacterial and viral illnesses are frequently accompanied by a transient loss of appetite, which is expected to self-resolve unless there is also growth faltering, which indicates severe sickness.^30^ Therefore, special attention must be paid to male, previously ill infants, infant growth trajectory, maternal mental health, type of feeds and frequency of feeding during each infant clinic visit.

## Limitations

One of the limitations of this study was the loss to follow-up of infants at the Maternal and Child Health (MCH) clinic. Additionally, the exclusion of small and sick babies from the study at birth may have artificially lowered the infant growth faltering rate in the cohort. The authors acknowledge the various definitions of infant growth faltering used in other studies, highlighting the absence of global consensus on standard growth definitions for infants below the age of 6 months.

## Strength

As a result of the logitudinal nature of the parent research, this article presented age-specific detailed growth statistics for infants under the age of six months, which are difficult to obtain in current literature.

## Conclusion ad Recommendations

Close attention must be paid to male infants, previously ill infants, infant growth trajectories, maternal mental health, types of feeds and frequency of feeding during the infant clinic visits. Infant cow milk feeding practices must be strongly discouraged. Absolutely, addressing lactational challenges, providing adequate support, regular and comprehensive infant anthropometry and assessment, are essential steps to promote exclusive breastfeeding (EBF) and ensure optimal infant growth and health. Lactating mothers require mental health support from health workers, family members and the community, in order to improve and sustain exclusive breastfeeding practices in the community. In rural areas, where access to healthcare services may be limited, strengthening maternal and child health (MCH) systems is crucial for providing the necessary support to breastfeeding mothers.

Implementing lactational support programs within rural MCH systems can involve various strategies, such as:

Education and Training: Healthcare workers should receive training on lactation management, including techniques for promoting successful breastfeeding, identifying and addressing common challenges, and providing support to mothers.

Community Health Workers: Utilizing community health workers (CHWs) who are well-trained in breastfeeding support can extend the reach of lactational support programs to remote areas. CHWs can provide education, counseling, and practical assistance to mothers in their communities.

Peer Support Groups: Establishing peer support groups for breastfeeding mothers can create a supportive network where mothers can share experiences, seek advice, and receive encouragement from one another. These groups can be facilitated by healthcare workers or community leaders.

Home Visits: Conducting home visits by trained healthcare workers or CHWs can allow for personalized support and assistance tailored to the individual needs of breastfeeding mothers and their infants.

Integration with Existing Programs: Lactational support programs should be integrated into existing MCH services to ensure sustainability and maximize reach. This integration can include incorporating breastfeeding education into antenatal and postnatal care visits and providing support through existing maternal and child health programs.

By strengthening lactational and mental support programs within rural MCH systems and work-places, mothers can receive the assistance they need to overcome lactational challenges and achieve full and optimal exclusive breastfeeding, ultimately promoting better infant growth and health outcomes in these communities.

## Author Contributions

The corresponding author (Dr Adenike Oluwakemi Ogah) conceived and designed the study, collected data and conducted data analysis, interpreted the results, and drafted the manuscript.

## Data Availability

All data produced in the present study are available upon reasonable request to the authors

## Acknowledgements

The author is extremely grateful to the participants involved in this study, to the staff of Gitwe Hospital and clinic in Rwanda and to the research team.

